# A new model to prioritize waiting lists for elective surgery under the COVID-19 pandemic pressure

**DOI:** 10.1101/2020.07.21.20157719

**Authors:** Roberto Valente, Stefano Di Domenico, Matteo Mascherini, Gregorio Santori, Francesco Papadia, Giovanni Orengo, Angelo Gratarola, Ferdinando Cafiero, Franco De Cian, Enzo Andorno, Giulia Buzzatti, Susan Campbell, Walter Locatelli, Marco Filauro, Marta Filauro, Marco Frascio, Carlo Introini, Franca Martelli, Guido Moscato, Giorgio Orsero, Giorgio Peretti, Paolo Pronzato, Edoardo Raposio, Mirella Rossi, Stefano Scabini, Nicola Solari, Carlo Terrone, Luca Timossi, Giovanni Ucci

## Abstract

The COVID-19 pandemic burdens non-covid elective surgical patients by reducing service capacity, forcing extreme selection of patients most in need. Our study assesses the SWALIS- 2020 model ability to prioritize access to surgery during the highest viral outbreak peaks.

A 2020 March - May feasibility-pilot study tested a software-aided, inter-hospital, multidisciplinary pathway. All specialties patients in the Genoa Surgical Departments referred for urgent elective patients were prioritized by a modified Surgical Waiting List InfoSystem (SWALIS) cumulative prioritization method (PAT-2020) based on waiting time and clinical urgency, in three subcategories: A1-15 days (certain rapid disease progression), A2-21 days (probable progression), and A3-30 days (potential progression). We have studied the model’s applicability and its ability to prioritize patients by monitoring waiting list and service performance. https://www.isrctn.com/ISRCTN11384058.

Following the feasibility study (N=55 patients), 240 referrals were evaluated in 4 weeks without major criticalities (M/F=73/167, Age=68.7 +/- 14.0). Waiting lists were prioritized and monitored. The SWALIS-2020 score (% of waited-against-maximum time) at operation was 88.7 +/- 45.2 at week 1 and then persistently over 100% (efficiency), over a controlled variation (equity), with a difference between A3 (153.29 +/- 103.52) vs. A1 (97.24 +/- 107.93) (p <0.001), and A3 vs. A2 (88.05 +/- 77.51) (p <0.001). 222 patients underwent surgery, without related complications or delayed/failed discharges.

The pathway has selected the very few patients with the greatest need, even with +30% capacity weekly modifications, managing active and backlog waiting lists. We are looking for collaboration for multi-center research.

## Full text

The reduction of routine surgery capacity during the COVID-19 outbreak triggers severe consequences on waiting lists, determining their impressive expansion with management costs^1^. The problem immediately burdens urgent and cancer patients, whose number of avoidable deaths indirectly due to COVID-19 is estimated close to that of SARS-Cov-2^2^. Planning and scheduling surgery becomes complex on clinical, ethical, and technical grounds. While several authors and professional associations have proposed clinical prioritization through urgency classifications^3^, pathways and data system models, specific tools are necessary to actually run priority-based-scheduling sustainably, in a usable and scalable fashion^4^. The Surgical Waiting List InfoSystem (SWALIS) has been previously proposed with such aims^5^. We report the pilot adoption of a new (SWALIS-2020) model to prioritize elective surgery during the COVID-19 pandemic (https://www.isrctn.com/ISRCTN11384058).

This is a 6-weeks (March - May 2020) feasibility-pilot cohort study testing a bespoke software-aided, inter-hospital, centralized multidisciplinary pathway serving all major elective urgent surgery from specialties in the 840,000-inhabitant Metropolitan area of Genoa. The pathway is based on centralized and MDT triage of referrals, further prioritized by the SWALIS-2020 model (Figure 1):

**Figure 1:**
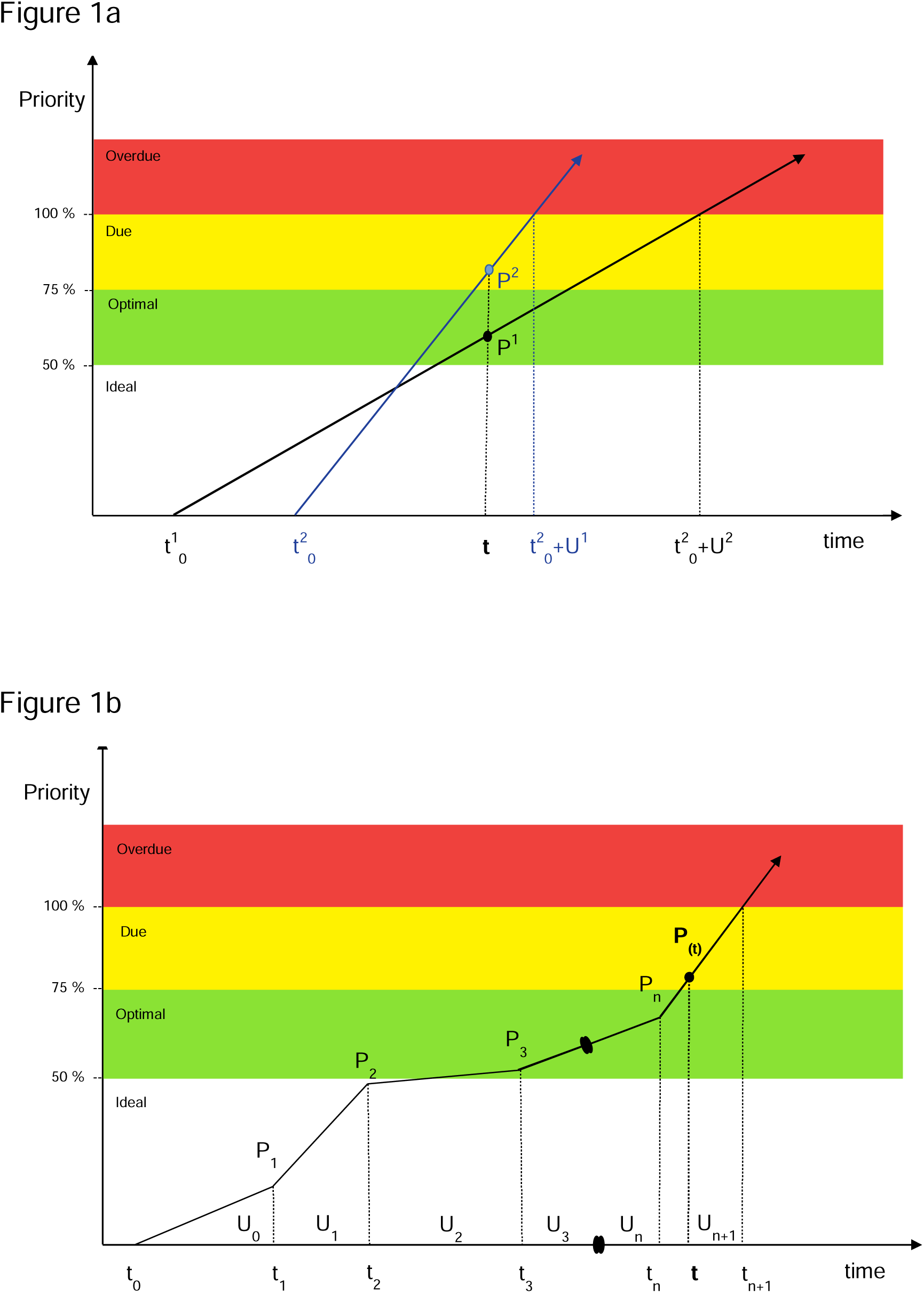
The prioritization method.

1. urgency categorization over maximum-waiting-time, defined by implicit clinical criteria: A1-15 days (certain rapid disease progression), A2-21 days (probable progression), A3-30 days (potential progression), B-60 days (no progression but severe symptoms), C-180 days (moderate symptoms), D-360 days (mild symptoms);
2. waiting list prioritization, real-time ordered by the SWALIS-2020 score (% of waited-against-maximum time) computed by a proportional, time-based linear cumulative method (PAT-2020, Figure 1);
3. theatre capacity planning, based on the prioritized demand;
4. flexible service-based priority-based scheduling.

We monitored pathway’s safety and efficacy by adverse events, drop-offs and completions, auditing its performance weekly by the SWALIS cross-sectional and retrospective waiting list indexes (dimensions and centrality), and by the SWALIS-2020 score at admission. Applicability was tested over pathway deviation events, number of postponements (prior to admission) and cancellations (on the day). Data was managed by live-running interface, code-developed on MS VBA™. Statistical analysis included use of Spearman’s rank for correlation, Mann-Whitney U test or one-way ANOVA with the Kruskal-Wallis rank-sum, the Dwass-Steel-Critchlow-Fligner or Loess tests, performed with the R software (version 3.6.3).

Following the 2-weeks feasibility phase (N=55 patients), 240 referrals were prioritized in 4 weeks without major pathway-related criticalities (M/F=73/167, Age=68.7±14.0). Waiting lists were monitored, and theatres fully allocated based on services’ prioritized demand. The SWALIS-2020 score at admission was 88.7±45.2 at week 1, then persistently over 100% (efficiency), over a controlled variation (equity), with a difference between A3 (153.29±103.52) vs. A1 (97.24±107.93) (p <0.001), and A3 vs. A2 (88.05±77.51) (p <0.001). Two-hundred-twenty-two patients eventually underwent surgery, without pathway-related complications or delayed/failed discharges.

While the different geographical areas are facing COVID-19 outbreak asynchronously, waiting list backlog will continue for months burdening hundreds of thousands of patients, and prioritization will long remain a major issue. The SWALIS-2020 model is designed for the broadest hospital acute care environment. It has smoothly selected and prioritized the very few patients with the greatest need, scheduling their access even with ∼30% capacity modifications weekly, managing active and backlog waiting lists in the same process. The heterogeneity of established practices in different services represents a challenge for waiting list pooling. However, the SWALIS-2020 model has passed the test, allowing effectiveness, efficiency, and equity. Our results encourage its wider adoption to prioritize surgery during the COVID-19 pandemic. We are looking for collaboration for further multi-center research.

--

No author declares competing interest. The study has not received external/*ad-hoc* funding. Prof. Angela Testi and Dr. Alessia Saverino gave insightful input in the manuscript. Ms. Marina Caldano input data. Prof. Susan Campbell was involved as a medical writer.

## Data Availability

The datasets generated during and/or analyzed during the current study are/will be available upon request from Mr Roberto Valente and Dr Stefano Di Domenico. The dataset is in an MS Access TM format, and can be anonymized to any level required, as allowed by the Liguria Ethics Committee. The dataset will be made available 3 months after study completion, for 12 months, extendable. Any sharing request will be submitted to the Liguria Ethics Committee for approval.

## 1a) The linear prioritization method (Pat 2007, SWALIS 2009)

The referring surgeon declares patient’s clock start date (t_0_) and clinical urgency category (U) based on the likelihood of quick deterioration, to the point where it may become an emergency, or on the level of symptoms, dysfunction or disabilities. Clinical urgency (U) is then associated to maximum waiting time from t_0._ In the SWALIS 2020 model U can assume six different values in days: U = {A1=15, A2=21, A3=30, B=60, C=180, D=360}.

Given on U and t_0_, and defining P(*t*_0_+U) = 1, the priority (P) at the time of prioritization P(t) is defined as follows:

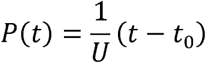

t^1^_0_ = patient 1 clock start date; U_1_ = patient 1 urgency category maximum allowed waiting time; t^1^_0_ = patient 2 clock start date; U_2_ = patient 2 urgency category maximum allowed waiting time; P^1^ = patient 1 priority at time of prioritization (t); P^2^ = patient 2 priority at time of prioritization (t).

## 1b) The cumulative linear prioritization method (Pat 2020, SWALIS 2020)

Clinical conditions can change during the waiting time (t_0_, t_1_, t_2_, … t_n_) affecting the patient’s urgency (U_0_, U_1_, U_2_, …U_n_). Priority can be calculated as summation, based on urgency variations:

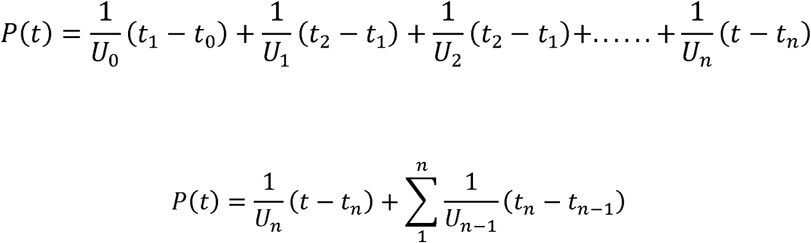

t_0_ = start waiting time; U_0_ = urgency for patient at starting time t_0_; t_n_ = updated urgency time; U_n_ = updated urgency for patient; t = time of prioritization.

The SWALIS 2020 prioritization method assumes four priority scores stages: “Ideal” (0-50%), color code white, “optimal” (51-75%) color code green, “due” (76-100%) color code yellow, “overdue” (>100%) color code red.

## References

1 Nepogodiev D, Bhangu A. Elective surgery cancellations due to the COVID□ 19 pandemic: global predictive modelling to inform surgical recovery plans. Br J Surg [Internet]. 2020 May 12 [cited 2020 Aug 9]; Available from: https://www.ncbi.nlm.nih.gov/pmc/articles/PMC7272903/

2 Maringe C, Spicer J, Morris M, Purushotham A, Nolte E, Sullivan R, et al. The impact of the COVID-19 pandemic on cancer deaths due to delays in diagnosis in England, UK: a national, population-based, modelling study. The Lancet Oncology. 2020 Aug; 21: 1023–1034.

3 Coronavirus □» Clinical guide for the management of essential cancer surgery for adults during the coronavirus pandemic [Internet]. [cited 2020 Apr 29]. Available from: https://www.england.nhs.uk/coronavirus/publication/clinical-guide-for-the-management-of-essential-cancer-surgery-for-adults-during-the-coronavirus-pandemic/

4 Déry J, Ruiz A, Routhier F, Gagnon M-P, Côté A, Ait-Kadi D, et al. Patient prioritization tools and their effectiveness in non-emergency healthcare services: a systematic review protocol. Systematic Reviews. 2019 Mar 30; 8: 78.

5 Valente R, Testi A, Tanfani E, Fato M, Porro I, Santo M, et al. A model to prioritize access to elective surgery on the basis of clinical urgency and waiting time. BMC Health Services Research. 2009 Jan 1; 9: 1.

